# Impact on Mental Health of students due to restriction caused by COVID-19 pandemic: Cross-sectional study

**DOI:** 10.1101/2021.02.07.21250695

**Authors:** Amar Prashad Chaudhary, Narayan Sah Sonar, Jamuna TR, Moumita Banerjee, Shailesh Yadav

## Abstract

**Background:** The aim of the study was to investigate fear, depression and anxiety symptoms among students of India due to COVID-19 pandemic and its restriction.

**Method:** The cross-sectional web-based research was conducted between mid-November and mid-December 2020 with the objective of understanding the psychological and behavioral consequences of the COVID-19 pandemic effect on students due to the constraint of forced control. The questionnaire included a) socio-demographic questions and b) psychometric scales evaluating the psychological and behavioral impact caused by COVID-19 pandemic and restrictions.

**Results:** Total number of 324 students participated in this study in which 44.4% were male and 55.6% were female. Fear of COVID-19 scale showed 68.8% of students had high fear, 24.4% had moderate to severe depression and 51.5% had moderate to severe anxiety. The correlation of fear of COVID-19 scale (FCV-19s) with Generalized Anxiety scale (GAD-7) and brief patient health questionnaire scale (PHQ-9) was found to be 0.492 and 0.474 respectively.

**Conclusion:** This research concludes that there is a very strong fear of COVID-19 among students, along with anxiety and depression symptoms. This study also concludes that the fear of the COVID-19 scale has strong positive correlation with the anxiety (GAD-7) and depression (PHQ-9) scales.

## Introduction

Communicable diseases such as herpes and legionnaire disease in the 1970s, HIV, Ebola, SARS, and now COVID-19 continue to be devastating, increasing pressure on people throughout the world (1). “The COVID-19 pandemic has rekindled the 21st century “viral scare” following the “microbe panic” of the 20th century. Public health acts like quarantine, physical distancing, use of face masks in public places and hand hygiene are executed globally to reduce the mushrooming of infection. Though these measures are effective to mitigate the pandemic, it has an adverse effect on the mental health of the people (2). Additionally, the quick transmission of coronavirus, increased death rate, absence of effective treatments and immunization, myth and misinformation have led to the increased fear, anxiety and depression among the students.

A recent study showed that feelings of anxiety and depressive symptoms, distress and sleep problems were typical signs of the COVID-19 pandemic (3). One of several important reasons for increased anxiety and depression is the threat of COVID-19 and, more specifically, the fear of becoming infected (4). Anger, exhaustion, psychological anguish, panic and confusion are the predominant psychological symptoms observed during COVID-19 pandemic is related to financial difficulties, annoyance and discomfort, loss of resources and ineffective coordination (5).

However, intensive fear may lead to decreased responsiveness and diminished mental well-being. Scientific reviews and analyses concentrate on identifying the various effects of COVID-19 not only on physical wellbeing, but also on mental health. The primary purpose of this research was therefore to explore students’ psychological responses, i.e. fear of COVID-19, anxiety and depression during the COVID-19 pandemic in India, and to also clarify causes of something which amplify the magnitude of COVID-19’s psychological effect. The second goal was to examine concurrent validity of Fear of COVID-19 scale (FCV-19s) with The Brief Patient health Questionnaire depression scale (PHQ-9) and Generalized Anxiety Disorder-7 scale (GAD-7) respectively.

## Material and method

### Study design & Study period

The cross-sectional web based study was carried out between mid-November to mid-December 2020 with the objective of understanding the psychological and behavioral impact caused on students due to restriction forced (physical distancing, use of face mask, online study, avoid gathering and going to public places, etc.) to control the COVID-19 pandemic effect. The survey questions and scale were selected on the basis of the available literature, author’s experience, and professor and clinician experience about the pandemic.

### Study site

For this study, students of Mallige college of Pharmacy in Bangalore, India are selected with access to the internet and Social media.

### Measures

The study included a) socio-demographic questions like Age, Gender, Degree Enrolled and Any of family members got infected with COVID-19, b) psychometric scales assessing the psychological and behavioral impact caused due to COVID-19 restrictions like fear, depression and anxiety. The scales are

#### Fear of COVID-19 scale (FCV-19s)

a self-report, unidimensional, reliable and valid scale developed recently to understand the fear of COVID-19 caused due to this pandemic. This scale consists of seven items which tries to explain the fear of COVID-19. The responses are recorded in 5 Likert scale ranging from 1 to 5 points (Strongly Agree =5, Agree=4, Neutral=3, Disagree=2, Strongly Disagree=1). The total Score ranges from 7 to 35. The higher the score is, the greater is the fear of COVID-19 among participants. The initial development of the scale has shown strong reliability of Cronbach alpha of 0.88 among the Iranian population (6). The cut off score for this scale is 19, the participant having score of 19 or more is considered to have high fear of COVID-19 and participant having score less than 19 is considered to have low fear of COVID-19 (7).

#### The Brief Patient health Questionnaire depression scale (PHQ-9)

is self-report, 9 item scale which is used to diagnose the major depression and subthreshold depression. The total score ranges from 0-27 which helps in interpretation of severity of depression. The responses of the participant are recorded in 4 points Likert Scale (not at all=0, several days=1, more than half the days=2, nearly every day=3). The cutoff score of 10 and above signifies moderate to severe depression with significant clinical concern whereas below score of 10 signifies minimal to mild depression (cutoff score: 0-4= minimal depression, 5-9=mild depression, 10-14=moderate depression, 15-19=moderately severe depression, 20-27= severe depression). The current study applied English for India version of PHQ-9 (8).

#### Generalized Anxiety Disorder-7 scale (GAD-7)]

is a self-report scale which was developed for initially diagnosing the Generalized Anxiety Disorders. The scale consists of 7 items for which the responses of the participant was recorded in 4 points Likert Scale (not at all=0, several days=1, more than half the days=2, nearly every day=3). The score of the participant ranges from 0 to 21. The threshold score of 10 has 89% sensitivity and 82% specificity for GAD (cut off score: 0-5= mild, 6-10= moderate, 11-15= moderately severe, 16-21= severe). The current study applied English version of GAD-7 (9).

### Sample size

The survey was completed using the Rasoft sample size calculator for capturing the appropriate sample size (10). The minimum of 306 samples were required for 95% confidence interval, 5% margin of error for the population distribution of 1500 students at 50% response distribution. Total of 324 students have taken part in this online study.

### Criteria of study

All the students studying diploma, undergraduate, graduate and post-graduate were selected for the study.

### Distribution of Questionnaire

The questionnaire was designed using Google forms and was distributed to the students through the various social media platforms like Whatsapp, Facebook, Messenger, Telegram, etc. By filling in the Google form without time constraint, the students were invited to participate in the survey. Multiple responses and double submission were eliminated by Google feature limit to one submission.

### Ethical Consideration

The purpose of the study was explained to the participating students and, prior to participating in the study, they were requested to submit their voluntary consent. All the procedures accomplished for this study were to adhere to the declaration of Helsinki 1964 and its later amendment (11). The online research was conducted and stated that the Checklist for Reporting Results of Internet E-Surveys (CHERRIES) guidelines were strictly agreed (12).

### Statistical Analysis

All the data were recorded in Microsoft Excel (Microsoft Corporation) and was assessed for the accuracy of the data. The statistical analysis was completed by using IBM SPSS software, version 25 (IBM Corporation). Descriptive statistics was carried out to understand the characteristics of the data (mean, standard deviation, range, maximum value and minimum value). Statistically, to understand the concurrent validity of fear of COVID-19 scale (FCV-19s) with Patient Health Questionnaire-9 (PHQ-9) and Generalized Anxiety Disorder-7 scale (GAD-7) Pearson Correlation was used. To understand the effect of independent variables like Age, Gender, Degree enrolled and Any of family members got infected with COVID-19 on these scales linear regression was used.

## Result

### Socio-demographic Characteristic

The socio-demographic characteristic of the participant is summarized in Table1. Among the total respondents of 324, 144 (44.4%) were male and 180 (55.6%) were female. The male respondents are less in comparison with the female respondent. The majority of the participants fall in the age group between 18-21 (58.6%) and 22-25 (35.8%). Most of the participants were enrolled in bachelor’s degree (79%). Among 324, 37(11.4%) reported that someone from their family member got infected with COVID-19 which seems to be low while comparing the transmission of this virus in the urban population.

**Table 1:** Sociodemographic characteristic

| **Socio-demographic Characteristic** |  | **Students Distribution (%)** |
| --- | --- | --- |
| **Age (in years)** | 18-21 | 190 (58.6%) |
|  | 22-25 | 116 (35.8%) |
|  | 26-29 | 8 (2.5%) |
|  | 30 and above | 10 (3.1%) |
| **Gender** | Male | 144 (44.4%) |
|  | Female | 180 (55.6%) |
| **Degree Enrolled** | Diploma | 11 (3.4%) |
|  | Bachelor's Degree | 256 (79%) |
|  | Master's Degree | 53 (16.4%) |
|  | PHD | 4 (1.2%) |
| **Any of family members got infected with COVID-19** | Yes | 37 (11.4%) |
|  | No | 287 (88.6%) |
| **Total Sample** |  | 324 |

### Psychometric scales

Descriptive statistics were studied for all three psychometric scales to understand the characteristic of the data which is summarized in Table 2. The magnitude of COVID-19 fears, symptoms of depression and anxiety were graded according to their cutoff ranking, which is explained in Table 3. There seems to be significant fear of COVID-19 (68%) and moderate to severe anxiety (51.5%) among the respondents. Only 28.7% of the respondents showed moderate to severe depression.

**Table 2:**
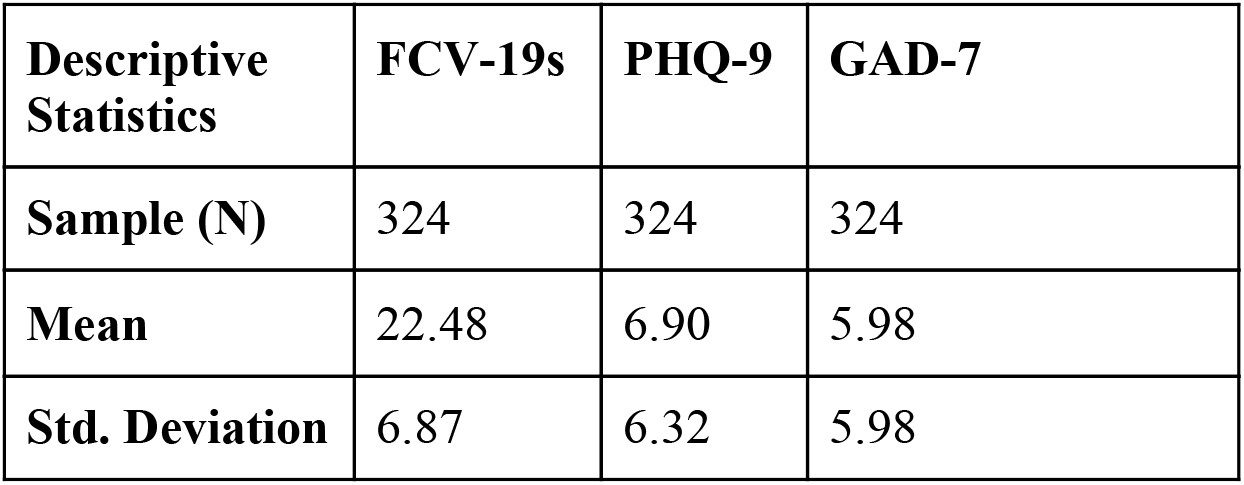
Descriptive Statistics of the sample according to the Psychometric Scale

**Table 3:**
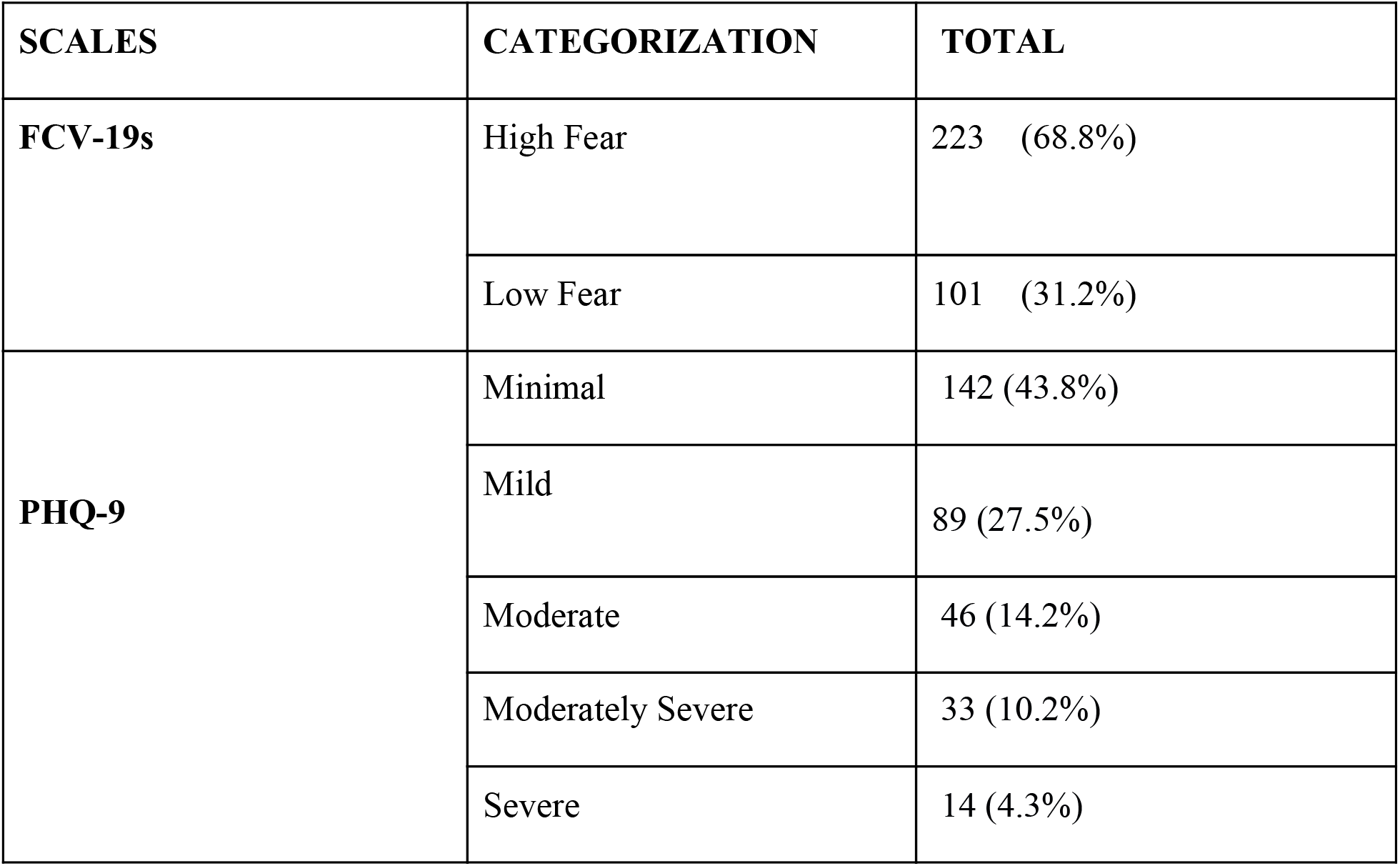
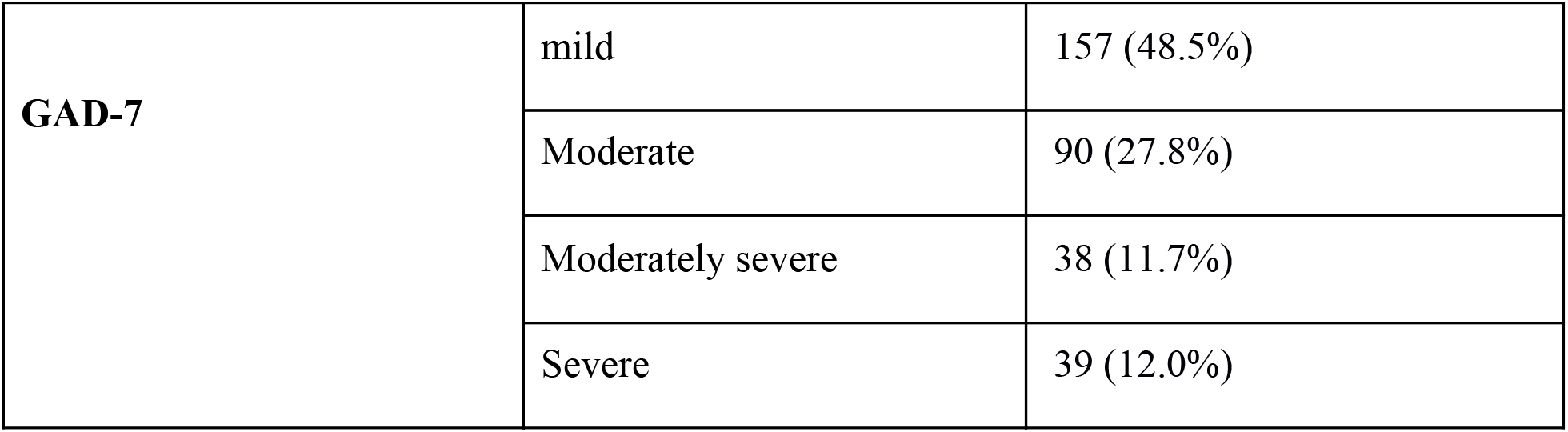
Categorization of students according to the scales cut off

There is strong association between the PHQ-9 scale and any family member who got infected with COVID-19 which is shown in table 4 but there seems to be not much impact of the independent variable like Age, Gender and Degree enrolled on any of the scale scores of participants. It explains that there is an increase in depressive symptoms when the number of COVID-19 infections increases in family.

**Table 4:** Correlation of key Variables using linear Regression with these scales

| **Scales** | **Age**<br>**p-value** | **Gender**<br>**p-value** | **Degree Enrolled**<br>**p-value** | **Any of family members**<br>**got infected with**<br>**COVID-19**<br>**p-value** |
| --- | --- | --- | --- | --- |
| **FCV-19s** | .049 | .542 | .276 | .468 |
| **PHQ-9** | .015 | .096 | .688 | .000** |
| **GAD-7** | .049 | .189 | .859 | .012 |

#### Fear of COVID-19 scale (FCV-19s)

In the survey, most of the respondents (63.7%) are found to be afraid of COVID-19, watching COVID-19 stories and reports on social media seems to have had a major effect on their mental health such that they become nervous or anxious. When they think about COVID-19, they couldn’t sleep properly due to fear of COVID-19 and about half of the respondents seem to feel uneasy thinking about it. Many people believe that their hands don’t become clammy or heart races when thinking of COVID-19.

#### The Brief Patient Health Questionnaire depression Scale (PHQ-9)

Many respondents (28%) think they have no motivation or enjoyment in doing stuff and feel down, sad or hopeless (21.9 percent). Approximately 20% of respondents appear to have difficulty falling asleep or sleeping for a long time or sleeping too much and find it difficult to focus on stuff (reading or watching television). About one fifth of the participants have shown the depressive symptoms of feeling tired and having less energy and feeling bad about themselves. Very few respondents have thoughts of better off dead or hurting themselves and poor appetite or overeating.

#### Generalized Anxiety Disorder (GAD-7)

About one-third of students feel that they become so restless, irritable and are afraid something awful happens to them several days. Around one fifth of the students agree that they feel anxious, worry too much, have difficulty relaxing half of day or nearly every day. Half of the students reported they don’t have such symptoms of anxiety.

### Concurrent Validation of Fear of COVID-19 Scale

There was found to be significant concurrent validity of fear of COVID-19 scale (FCV-19s) with anxiety (GAD-7) and depression (PHQ-9) scales (Table 5). The Pearson correlation of FCV-19s with PHQ-9 and GAD-7 was found to be 0.474 (p-value= 0.000) and 0.492 (p-value=0.000) respectively. This strong relationship helps to predict an increase in fear of COVID-19 will ultimately increase the anxiety and depressive symptoms in students. The fear of COVID-19 scale can give the overall idea regarding fear of COVID-19, depression and anxiety among the students.

**Table 5:** Concurrent validation of Fear of COVID-19 scale

| **FCV-19S Correlations with GAD-7 and PHQ-9** |  |  |
| --- | --- | --- |
| **FCV-19s**<br>**Sig. (2 tailed) (p-value)**<br>**N** | Pearson Correlation<br>324 |  |
| **GAD-7**<br>**Sig. (2 tailed) (p-value)**<br>**N** | Pearson Correlation<br>.000<br>324 | 0.492** |
| **PHQ-9**<br>**Sig. (2 tailed) (p-value)**<br>**N** | Pearson Correlation<br>.000<br>324 | 0.474** |

\*\*Correlation is significant at the 0.01 level (2-tailed)

## Discussion

### Sociodemographic

The survey was distributed by the most popular social networking platform in India via social media, including Facebook, WhatsApp and Instagram. The most common age of the volunteers in the survey was between 18-25 and between 18-21 (58.6%), 22-25 (35.8%), 26-29 (2.5%) and above 30 (3.1%) and participants were mainly from Bachelor’s degrees (79%). Compared to the guy, there are a greater number of female respondents statistically, but there is not much substantial difference. The gender distribution between males and females is around (44.4%) males and females (56.6%) respectively. In psychological and behavioral responses to the COVID-19 pandemic, the study conducted by parlapani et al. also showed higher numbers of female participants compared to male respondents (13). Women are found to have greater mental literacy and are more involved than men in engaging in health-related online studies (13,14).

### The Psychological Impact of COVID-19

Fear is considered a biologically “reflex” emotion, an automatic “basic” reaction to a specific external threat. There is a broad correlation between fear and anxiety, with an unknown, unclear or future risk being more associated with both. Via an evolutionary perspective, fear is related to risk-avoiding activities, while anxiety is linked to preparation. Adaptation and consciousness promote both emotional states. It may elicit behavioral regulation and promote self-protective responses, because fear is connected to self-perseverance (15,16).

The research conducted in Greece found that 35.7% displayed a higher degree of fear of COVID-19 (13). This study found that 68.7% of students have a greater degree of fear of COVID-19. The cross-sectional study found that one in every three participants had moderate to severe symptoms of anxiety during the initial phase of the pandemic in China (17,18). Only after the outburst of the pandemic, a cross-sectional analysis conducted in Shanghai and Wuhan showed that the incidence of mild to extreme anxiety increased 4-5 times (17). In Greece, a cross-sectional analysis to clarify the behavioral and psychological reactions to the COVID-19 pandemic found that 77.4% of participants had moderate to serious anxiety symptoms and 22.8% had moderate to severe depressive symptoms (13). The COVID-19 pandemic has been shown to cause increased anxiety, which is similar to other infectious diseases outburst. This study also found similar results, with more than 50% students have moderate to serious symptoms of anxiety and 28.7% students have moderate to severe depressive symptoms due to this pandemic and its restrictions.

### Concurrent Validation of Fear of COVID-19 Scale

Fear of COVID-19 scale is the recently developed tool to understand the fear caused by COVID-19 pandemic among the public. This scale has been validated with various established other psychometric scales which are used to understand anxiety, depression, etc. The Greek version of COVID-19 scale, have shown significant concurrent validity with GAD-7 scale (r=0.71, P-value <0.001) and moderate correlation with PHQ-9 Scale (r=0.47, P-value < 0.001) (7). The study conducted by Ahorsu et al. validated Fear of COVID-19 scale with hospital anxiety and depression scale (HAAD) had found a significant association (depression r = 0.425 and anxiety r = 0.511) (6). This study also found similar association with the well-established depression scale (PHQ-9) and anxiety scale (GAD-7) (depression r = 0.474, P-value < 0.001 and anxiety r = 0.492, P-value < 0.001).

## Conclusion

The psychological effect of COVID-19 in Indian students during the near pandemic year was analyzed in this study, and even the specific connections among fears, depression and anxiety of COVID-19. This research concludes that, along with anxiety and depression, there is a very strong fear of COVID-19 among students. This study also concludes that the fear of the COVID-19 scale and the GAD-7 and PHQ-9 scales have a strong positive correlation.

Comprehensively, although past experiences with the 2003 SARS epidemic has shown that infectious disease crises can have fast mental health effects, the long-term impact of COVID-19 needs to be assessed for the population of India. In order to mitigate fear and encourage healthier lifestyles during the pandemic, strategic public health interventions are needed.

## Study Limitations

Some of the constraints are included in this report. In this sample, there is an unequal distribution of respondents and it is a cross-sectional study so that casual intervention cannot be done. There are very few respondents from diploma students, so the survey result cannot generalize the student population as a whole. The questionnaire was self-administered and recorded in this review, so it is difficult to understand whether it was reasonably completed (i.e. social desirability bias and the answers do not show the reality). Since it is an internet-based survey, the study will not actively collect the responses of learners who are not linked to social media.

## Data Availability

this data in this study can be used to plan to improve mental health of students. this data can't be reproduced without the author approval.

## Acknowledgement

No funding has been granted for this study by any institution and organization. We are very thankful to the volunteers of this study and the college staff for making this study possible.

